# Disclosure of Gender Identity among Transgender Individuals to Healthcare Professionals in China: An Online Cross-sectional Study

**DOI:** 10.1101/2022.08.26.22279241

**Authors:** Shamen Susan Chauma, Chengsong Wan, Willa Dong, Xuezhen Fu, Joseph D Tucker, Gift Marley, Weiming Tang

**Affiliations:** School of Public Health, Southern Medical University, Guangdong Province, Guangzhou, 510515, China; Dermatology Hospital of Southern Medical University, Guangdong Province, Guangzhou, 510095, China; Gilling’s Global School of Public Health, the University of North Carolina at Chapel Hill, Chapel Hill, NC, 27599, USA; International Department of Nanjing N0.13 Middle School, Nanjing, China, 210000; University of North Carolina at Chapel Hill Project China, Guangzhou, 510095, China; Institute of Global Health and Infectious Diseases, the University of North Carolina at Chapel Hill, Chapel Hill, NC, 27599, USA

**Keywords:** China, Disclosure, Health Professionals, Health Services, Sexual Behaviors, Transgender Individuals

## Abstract

**Purpose:** Culture and stigma-relevant issues discourage transgender (TG) individuals in China from disclosing their gender identities. This limits their access to transgender competent health services. This study evaluates the factors associated with gender identity disclosure to health professionals among transgender individuals in China.

**Methods:** A cross-sectional study was conducted in nine cities across mainland China from December 2019 to June 2020 among transgender individuals. Participants completed questions covering socio-demographic information, HIV/STI testing, sexual behaviors, and access to medical and mental health services.

**Results:** Overall, 277 (85.5%) out of 324 transgender individuals were eligible to participate in the study. The mean age was 29±8 years old. Among them, 78% (192/277) had ever disclosed their gender identity to others, and 56% (155/277) had disclosed their gender identity to health professionals. 79.4% had ever tested for HIV (with an HIV prevalence of 9.1%), 47.3% had tested for other STIs, 42.6% had used hormone therapy, and 9.4% had undergone gender-affirming surgery. Results from the multivariable logistic regression demonstrated that compared to non-disclosers, disclosers were more likely to have tested for STIs (aOR=1.94, 95%CI: 1.12-3.39). Hormone intervention therapy (aOR=2.81, 95%CI: 1.56-5.05) and Pre-Exposure Prophylaxis (PrEP) use (aOR= 3.51, 95%CI: 1.12-10.97) were associated with gender identity disclosure to health professionals.

**Conclusions:** Low rates of gender identity disclosure to health professionals among transgender people may reflect fear of stigma and outing, suggesting the need for more trans-inclusive environments. More research is needed to understand the importance of disclosure in improving transgender health services.

**Summary:** A study among transgender individuals in nine cities across mainland China found that gender identity disclosers to health care professionals were more likely to test for STIs than nondisclosures.

## Introduction

Transgender is an umbrella term to describe people whose gender identity or expression differs from the gender assigned at birth based on sex. These individuals are a diverse group of people, not based on gender identity only but also based on age, ethnicity, socio-economic status, and their engagement in HIV risk activities. ^2, 3^ Given the stigma associated with being transgender in many settings, gender identity disclosure to health professionals is challenging for transgender individuals. ^1, 4^ Transgender people are vulnerable to various adverse health outcomes five and have a higher prevalence of HIV and other STIs mental health distress, gender minority stress, substance use, and abuse.^7^Toimprove the health and well-being of transgender people, it is necessary to provide transgender competent services. Disclosure should be encouraged because it increases the likelihood of higher quality services.^4, 8^

Understanding transgender health entails becoming aware of the health risks that transgender individuals face. Coming out to health professionals as transgender is fraught with emotional and physical dangers. Although disclosure creates an opportunity to discuss specific issues and concerns,^9^ it could inadvertently lead to discrimination and unequal treatment by health professionals. According to a 2011 study conducted in China, approximately 49% of transgender patients in the hospital setting were denied treatment, disrespected, and physically attacked. As a result, a significant proportion of Lesbians, Gays, Bisexuals, Transgender, and Queer (LGBTQ) patients deliberately conceal their gender identity from health professionals. However, failure to disclose gender identity could have detrimental implications for comprehensive transgender health services, including sexual health and others.^10^ Furthermore, institutional barriers to care, such as shared hospital rooms, shortage of experienced trans-gender health personnel, and lack of specialized care, ^11, 12^have increased the reluctance to disclose.

With over 1/3 of transgender individuals resisting disclosure, possible effects such as low health care seeking, low health support, severe stigma, substance use, and poor mental health are widespread.^13^According to one LGBTQ report, only 35% of respondents disclosed their identity to health professionals.^14^Another study^15^ revealed that sexual activity, HIV and STIs monitoring, and transmission of transgender individuals were negatively affected by the low level of disclosure, but no study in China has evaluated this situation.

Globally, an estimated 0.3-0.5% of the population is identified as transgender ^16^, and China is identified as one country in Asia with a strong transgender community. An estimated 4 million transgender people live in China, of which 4-17% live with HIV.^17^Owing to the potential social and cultural pressures of the Chinese society towards transgender individuals, they tend to conceal their gender identity.^18-20^ Despite having high HIV prevalence, their use of HIV prevention services such as PrEP is low.^6, 21^ One of the United Nations Program on HIV/AIDS (UNAIDS)global targets is to attain 95-95-95 by 2030, and to achieve this feat, key population such as transgender individuals must be included in HIV combined care. Gender identity disclosure information is vital as it helps incorporate the needs of TG individuals in HIV prevention strategies.

However, the lack of focus on trans issues in Chinese medical training provides a unique environment to conduct this study in China. Most physicians also do not ask patients about gender identity as little is known about their healthcare interactions.^22^ In addition, no study in China has examined the relationship between gender identity disclosure to health professionals and healthcare service utilization, with a particular emphasis on especially among transgender people. Many studies that reported on health care service utilization generalized the LGBTQ community.

This study is of public health importance because assessing disclosure rate and health care utilization among transgender is a reliable approach to combating the global HIV burden. Therefore, this study aims to understand the status and factors associated with gender disclosure among Chinese transgender individuals and examine the relationship between gender identity disclosure to health professionals and uptake of healthcare services.

## Methodology

### Ethical statement

This study was approved by the Institutional Review Boards (IRB) #18-3215 of the UNC at Chapel Hill and the Dermatology Hospital of Southern Medical University before the survey’s launch.

### Study design and sampling methods

The study was organized by the University of North Carolina (UNC) Project-China. An online survey was conducted among transgender individuals in Guangzhou, Nanning, Kunming, Nanjing, Guiyang, Shenyang, Jinan, Qingdao, and Wuhan. These nine cities in China were chosen from demographically diverse areas with local Community-based organizations (CBOs) that provide services specifically to transgender people, including linkage to HIV and STI services.

Potential participants who visited or were served by the partnering CBOs were informed about the study and asked if they were willing to participate. During outreach activities, the staff of the CBOs introduced the study to potential participants. Potential participants were told about the study privately at the partnering CBO to protect participant privacy. The study inclusion criteria were: 18 years old or older, could speak Mandarin Chinese; could provide consent; assigned male gender at birth but currently does not identify as a man. Eligible participants provided informed consent by checking a box indicating their agreement to participate in the study voluntarily. The 30 minutes surveys were completed at the CBO offices on a laptop, tablet, or smartphone. The study staff was available to answer questions or for technical assistance.

### Measures

Eligible participants recruited were required to visit specific CBOs to complete the study survey. However, this method was switched to an online survey approach later due to the Corona Virus Disease 2019(COVID-19) pandemic. The anonymous survey collected information on socio-demographics, gender disclosure, and sexual behaviors. Socio-demographic information included marital status (never married or married, divorced or separated or widowed), level of education (high school or below, including technical secondary school, associate, college, graduate or above), residence (one of the cities understudy), and annual income (less thanUSD3000, USD3001–6000, USD6001–10,000 or more than USD10, 000).The survey also collected information on healthcare services which included hormone intervention use (yes or no), gender-affirming surgery(yes or no), and PrEP use(yes or no). In addition, participants were asked if they had ever tested for HIV (yes or no), tested for other STIs like gonorrhea, Chlamydia, and syphilis (yes or no), and engaged in condomless sex (yes or no) in their lifetime. Finally, participants were also asked questions about their experience in society with family and friends. Regarding gender identity disclosure, participants were asked if they had ever mentioned their gender identity to others (yes or no) and if they had ever disclosed their gender identity to their healthcare providers (yes or no).

### Statistical analysis

We segregated the socio-demographic and gender disclosure details for descriptive analysis by whether participants had disclosed their gender identity to others and healthcare professionals. Multivariable logistic regression models were used to assess correlations with gender identity disclosure among the study participants. Factors that were adjusted in the multivariable analyses included residence (Guangzhou, Nanning, Kunming, Nanjing, Guiyang, Shenyang, Jinan, Qingdao, and Wuhan), educational level (high school or below, including technical secondary school, associate, college, graduate and above), monthly income (less thanUSD3000, USD3001–6000, USD6001–10,000, or more than USD10,000), and marital status (never married, married, separated, widowed).

All data analyses were completed using IBM SPSS statistics 26.

## Results

A total of 324people started the survey, and a total of 277 (85.5%) participants met the inclusion criteria and completed the online survey. About 44%of participants had not revealed their gender identity to health care providers, and 22% had not revealed their gender identity to anyone.

### Socio-Demographic characteristics of participants

The mean age of the 277 included participants was 29.8± 7.9 years old. Overall, 45.5% (*n* = 126) had completed high school or less, 3.6% (*n* = 10) were married, 88% (*n=*243) had never married, and 8.3 % (*n=*23) were separated or divorced. In addition, 46.6% (*n* = 129) of the participants were currently receiving an annual income of more than USD 3000 and less than USD 6000, and the minority of the participants (26.4%, *n* = 73) had an annual income of more than USD6, 000 (Table 1).

### Behavioral characteristics

In total, 78 % of participants (*n* = 192) had disclosed their gender identity to others (including family and friends), and 56% of participants (*n* = 155/277) had ever disclosed their gender identity to a healthcare provider. 53.3 % (*n*=48) of participants had a stable sexual partner in the past three months, 85.5% (*n*=53) reported condomless sex, and 14.5% (*n*=9) consistently used condoms with their partner in the last three months. About 44.8 % (*n*=69) reported being involved in lifetime sex work, 41.9 % (*n*=65) reported previous tobacco use, 66.5 % (*n*=103) reported alcohol use, and 14.2 % (*n*=22) had ever used recreational drugs. A 49.7 % *(n*=77) of participants reported difficulties accessing medical treatment due to their gender identity, while 55.5% *(n*=86) reported getting poor medical services due to their gender identity. 47.1 % (*n*=73) felt that the society is never fair to them, 58.1% (*n*=90) felt rejected by the society due to their gender identity, and 38.7 % (*n*=60) reported having emotional help and support from friends and family (Supplementary Table S1)

Among all the participants, 79.4% (*n*= 220/277) had ever tested for HIV [with HIV prevalence of 9.1% (*n*=20/220)] and 47.3 % (*n*=131/220) had ever tested for other STIs. The majority of participants (83.9%, n = 130/277) who had tested for HIV and 54.2 % (*n*=84/131) of participants who had tested for STIs had disclosed their gender identity to health professionals. Furthermore, 54.2% (*n*=84) of participants reported using hormone intervention therapy, and 11.6% (*n*=18) who had undergone gender-affirming surgery had disclosed their gender identity to health professionals. Few participants (12.3 %, *n*=19) who had disclosed their gender identity to health care providers reported ever using PrEP (Table 2)

### Factors associated with overall gender identity disclosure

Compared to non-disclosers, participants who had disclosed their gender identity were more likely to have tested for STIs with aOR of 1.82 (95%CI: 0.98-3.39). The likelihood of gender identity disclosure was two times higher among transgender individuals who reported previous use of alcohol with aOR =2.44 (95%CI: 1.31-4.55) and was low among transgender individuals who used recreational drugs aOR=0.37 (95%CI: 0.12-1.12). Participants who had ever used hormone intervention therapy were more likely to have disclosed their gender identity with aOR of 1.86 (95%CI: 0.93-3.74) than those who had never used hormone therapy (Supplementary Table S2).

### Factors associated with gender identity disclosure to healthcare providers

Factors associated with disclosure of gender identity to healthcare providers were also assessed in our study. The likelihood of gender identity disclosure to healthcare providers was greater among transgender who had ever tested for HIV (aOR =1.72, 95%CI: 0.87-3.39) and STIs (aOR=1.94, 95%CI: 1.12-3.39). Also, PrEP use (aOR= 3.51, 95%CI: 1.12-10.97) and use of hormone intervention therapy (aOR = 2.81, 95%CI: 1.56-5.05) were associated with a higher likelihood of gender identity disclosure to health professionals (Table 3).

## Discussion

Understanding the current status of gender identity disclosure is essential in providing tailored services to transpeople. This study extends the existing literature by recruiting participants from various demographic and economic regions of nine cities in China using an online recruitment process. We found that 44% of transgender persons have not disclosed their gender identity to health professionals, and 22% had not revealed their gender identity to anyone. Gender identity disclosure was linked to HIV and STI testing services, PrEP use, and the use of hormone intervention therapy.

We observed low rates of gender identity disclosure to health professionals among transgender people. Similar studies in Canada and Chicago found that gender identity disclosure to primary care providers was equally as challenging as disclosing to others with over a third of transgender adults refusing to do so, which presented a barrier to obtaining quality health care.^23, 24^ Many factors contribute to the low rate of gender identity disclosure to health professionals. Studies have documented the reasons for non-disclosure as the providers’ failure to ask about identity, internalized stigma, and the assumption that health and gender identity are unrelated. Furthermore, when participants revealed their identities, they received negative reactions.^25^

Another study conducted in China found that approximately 8.0% of Chinese LGBTQ people experienced negative treatment (including discrimination and verbal abuse by medical staff) in medical care settings and received poor medical care^.26^ Poor pathways, unmet primary health needs among transgender populations, insufficient training and knowledge on trans-specific health care by providers, gender-based discrimination within the health care system, and poor stakeholder participation in targeted interventions development may have contributed to the low rate of gender identity disclosure among transgender people in China. ^27^ The difficulty in disclosing gender identity can be reduced by a strong therapeutic relationship between care providers and trans individuals and structural interventions to make the local environment more welcoming for trans people in specific clinical settings.

PrEP use was also positively associated with gender identity disclosure, but the rate of PrEP use was extremely low among transgender people in China. PrEP is an intervention to support key populations, including transgender people, with HIV risk behaviors and reduce their risk of HIV infection. However, studies on PrEP uptake among transgender people have yielded disappointing results, with a lack of efficacy attributed to a low adherence rate. In 2019, a systematic review on HIV-related care for trans-people found that there are limited data on PrEP use among trans people and that access and consistent usage remain poor among the population globally.^28^ A national online survey conducted in the United States found that only 48 transgender individuals reported actual PrEP use out of 1800 transgender persons who had access to it. ^29^ Another online study found that among the 84% of participants who had heard of PrEP, only 33% reported ever using PrEP, and only 22% reported use at the study time.^30^ In addition, a study in New York City found that only 13.3% of trans individuals had ever used PrEP.^31^However, according to a Jamaica study, increasing PrEP awareness among transgender people who engage in sex work is critical because sex work is linked to PrEP awareness and acceptability. ^32^A New York study found that unwillingness to use PrEP was associated with concerns about side effects. ^33^

Other reasons for the low PrEP use among transgender people worldwide include unwillingness to pay for it, low positive expectations of its efficacy, and no experience with HIV testing.^34^ Increased PrEP uptake among the transgender could be achieved by publicizing information on PrEP, emphasizing the sexual health benefits of using PrEP, improving patient-provider communication, expanding the available HIV prevention strategies, and conducting more research studies to evaluate PrEP in transgender individuals. ^28,35^Patients should be introduced to PrEP while seeking general health services. The specific needs of transgender people should be considered to develop inclusive patient-centered, and LGBT-friendly services.^10^ This will increase self-assurance among transgender persons and encourage them to disclose their gender identity to obtain holistic healthcare.

Our study findings showed that gender identity disclosure to health professionals is also linked to HIV and STI testing. This finding aligns with a study carried out in Thailand among transgender women.^36^Another study conducted in China found similar associations where HIV and STI testing were associated with further discussions of HIV risks between people engaged in same-sex activities and their doctors. ^37^ The study showed that China had a low percentage (34.6%) of transgender people who had tested for HIV in the past. The results also suggested that the fear of being labeled as an LGBTQ person deterred them from having tested for HIV and other STIs. Many transgender individuals also engage in sex work, which increases their risk of HIV and STI infection. As a result, health professionals aware of their patients’ sexual risk behaviors are more likely to inquire about their gender identity and offer STI and HIV prevention services.

Literature has shown that transgender people who disclose their gender identity to health professionals are more likely to get basic health checks, including HIV and STI services. Therefore, expanding HIV and STI testing services could positively influence gender identity disclosure to health professionals and should be considered during project implementations. Furthermore, health services should be tailored to bridge the gaps in access to healthcare for people afraid of being stigmatized for going to sexual health clinics or being identified as transgender. Given the high burden of HIV among transgender individuals, improving gender disclosure could help increase HIV and STI testing uptake and thus aid in the fight against the global HIV pandemic. This can be achieved by having transgender-friendly and non-stigmatizing health services.

According to the findings of this study, 58.1% of participants who had disclosed their gender identity experienced social and family rejection, while 38.7% reported receiving emotional support and help from friends and family. Family and social rejection based on gender identity is an interpersonal stressor that can harm the mental health of transgender people. To avoid discrimination, stigma, and related problems, many transgender people hide their gender identity from health professionals, family, and friends. According to a review study about mental health among TG, a higher percentage of transgender people who received mental health services (42.8 %) would not be hesitant to reveal their gender identity, compared to 11% who would be hesitant. ^12, 38^ Mental health concerns, on the other hand, can discourage transgender people from pursuing gender-specific health interventions such as gender-affirming surgery and hormone therapy, both of which typically require gender disclosure to health professionals.

According to one systematic review, gender-affirming therapy seems to have overwhelming beneficial psychological results. Research supports this intervention to alleviate anxiety and depressive symptoms, lower social distress, and improves the quality of life and self-esteem of transgender individuals.^39^ Even though overcoming socio-cultural barriers to promoting gender disclosure remains a challenge in China and other LMICs, community campaigns, family engagement, and social support from family, friends, and healthcare providers could help address mental stress in transgender people.^40^ Because a supportive relationship with family and health professionals fosters trust and a sense of belonging that decreases feelings of shame and could help facilitate willingness to disclose and use gender-specific health care among transgender people.^41^

### Implication

The results of the study have several policies and research implications. The Chinese transgender people have a low rate of gender disclosure to health care providers due to various factors. From the policy perspective, policymakers should collaborate with CBOs and other stakeholders to increase the competence of STD doctors in serving transpatients. This would help meet transgender expectations while receiving care and thus promote gender disclosure, as a relaxed and therapeutic environment significantly impacts gender identity disclosure.^42^

Also, policymakers should consider developing operational guidelines for healthcare providers serving transgender individuals to conveniently allow transgender people to receive gender-affirming services. Moreover, policymakers should consider the kind of health professionals to whom transgender individuals are likely to disclose their identity and provider behaviors that may make disclosure easier.

From the research perspective, future research should focus on defining barriers and facilitators to gender identity disclosure, how disclosure impacts healthcare decision-making among transgender people and the effect of positive and negative disclosure experiences on future health-seeking behaviors. Also, researchers should look at transgender people’s pre-and post-gender re-assignment intervention needs; and health professionals should be able to offer gender-affirming treatments that are non-discriminatory, comprehensive, and of high quality. More research is required to explore pathways that can guide action, like identifying marginalized groups at higher risk of poor health and agreeing on potential invention goals like peer support and community participation.

### Limitations

Our study has several limitations. First, this was a cross-sectional study, and hence no causal relationships can be inferred from our results. Second, recruited participants were predominantly young and well-educated transgender individuals in an online sample. This sample probably excluded older transgender persons who were more likely to be less educated and technologically challenged. Given a longer exposure to social stigma, this population might be less likely to disclose their gender identity. Third, there may have been a selection bias as participants exempted due to incomplete surveys may have had different socio-demographic features and behaviors. Fourth, “healthcare professional” was a generic term in this study that included various healthcare providers, including HIV clinics, hospitals, testing staff, general medical practitioners, and more. Since the roles of these care providers vary, our results cannot be generalized with any certainty. Also, we may have ignored important details by considering the different types of providers as one general category, and the approximate connections may be distorted. Future studies should resolve this problem with subgroup analyses to provide more information on the subject. Lastly, the disclosure question was not desegregated by physician elicited disclosure versus self-prompted disclosure.

## Conclusion

This study found that disclosure of gender identity is associated with HIV and other STI testing, PrEP use, and hormone intervention uses. To promote gender identity disclosure among transgender people, policymakers, researchers, and the transgender community should work together to create a more inclusive and supportive environment where PrEP and other health services are easily accessible. Future longitudinal research on how gender identity disclosure affects transgender sexual behaviors is needed to provide targeted interventions. Promoting gender identity disclosure may be part of a comprehensive HIV intervention tailored to meet the needs of transgender people in China and other Low- and Middle-Income Countries.

## Supporting information

References

## Data Availability

All data produced in the present study are available upon reasonable request to the authors

## Author Contributions

WT, WD, & JDT conceived and designed the study; SC, WC, WT & XF acquired, screened, analyzed, and interpreted data for the work; and SC & WT drafted the manuscript. WT, WC, JDT, WD, GM & XF critically revised it for important intellectual content. All authors approved the final manuscript version for publication. They agreed to be accountable for all aspects of the work in ensuring that questions related to the accuracy or integrity of any part of the work were appropriately investigated and resolved. The corresponding author attests that all listed authors meet authorship criteria and that no others that meet the criteria have been omitted.

## Acknowledgment

We thank all the participants and funders who contributed to the successful completion of this study. In addition, we appreciate YongJie Sha and LingLing for serving as scientific advisors during this study.

## Funding statement

This study received support from internal institute funding from Zhuhai Center for Disease Prevention and Control. This work was also supported by the National Nature Science Foundation of China (81703282 and 81903371), the National Institutes of Health (NIAID 1R01AI114310-01, NIAID K24AI143471), NIMH (R34MH109359 and R34MH119963), and National Science and Technology Major Project (2018ZX10101-001-001-003). Willa Dong was funded by a Fulbright-Hays Doctoral Dissertation Research Abroad fellowship; the Graduate Tuition Incentive Scholarship and Dissertation Completion Fellowship, The Graduate School, The University of North Carolina at Chapel Hill; the STD/HIV T32 Pre-doctoral Training Program from the National Institute of Allergy and Infectious Diseases (grant number 5T32AI007001-42); The funders did not participate in the process of study design, data collection, and analysis, decision to publish, or preparation of the manuscript. The authors thank all people who contributed.

## Disclosure statement

The authors report no competing interests.

## Notes

### Competing Interest Statement

The authors have declared no competing interest.

