## Supplementary material for "Disclosure of Gender Identity among Transgender Individuals to Healthcare Professionals in China: An Online Cross-sectional Study": Tables

| Table 1:Socio- Demographic Characteristics of 277 Chinese Transgender Individuals, 2020. | | | | | | | | | | | | | | | | | |
| --- | --- | --- | --- | --- | --- | --- | --- | --- | --- | --- | --- | --- | --- | --- | --- | --- | --- |
| **Variables** | **To Others** | | | | | **To Healthcare Professionals** | | | | | | | | | **Overall (N =277)** | | |
|  | **Non-disclosers(n=61)** | | **Disclosers(n=192)** | | | **Non-disclosers (n=122)** | | | | **Disclosers (n=155)** | | | | |  |  |  |
|  | N | Percent (95%CI) | N | | Percent (95%CI) | N | | Percent (95%CI) | | N | | | Percent (95%CI) | | N | Percent (95%CI) | |
| **Age** | | | | | | | | | | | | | | | | | |
| <20 | 3 | 4.9 (1~13.7) | 9 | | 4.7 (2$.$2~8.7) | | 7 | | 6.4(2.6~12.8) | | 5 | | 3.5 (1.1~7.9) | | 12 | 4.7 (2.5~8.1) | |
| 20-29 | 31 | 50.8 (37.7~63.9) | 99 | | 51.6  (44.3~58.8) | | 58 | | 53.2(43.4~62.8) | | 72 | | 50 (41.6~58.4) | | 130 | 51.4(45~57.7) | |
| >29 | 27 | 44.3 (31.5~57.6) | 84 | | 43.8 (36.6~51.1) | | 44 | | 40.4(31.1~50.2) | | 67 | | 46.5 (38.2~55) | | 111 | 43.9(37.7~50.2) | |
| **Residence** | | | | | | | | | | | | | | | | | |
| Current city | 47 | 67.1 (54.9~77.9) | 110 | | 53.1 (46.1~60.1) | | 82 | 67.2 (58.1~75.4) | | | 75 | | | 48.4 (40.3~56.5) | 157 | | 56.7 (50.6~62.6) |
| Other city in the province | 6 | 8.6 (3.2~17.7) | 24 | | 11.6 (7.6~16.8) | | 11 | 9 (4.6~15.6) | | | 19 | | | 12.3 (7.5~18.5) | 30 | | 10.8 (7.4~15.1) |
| Outside the province | 17 | 24.3 (14.8~36) | 73 | | 35.3 (28.8~42.2) | | 29 | 23.8 (16.5~32.3) | | | 61 | | | 39.4 (31.6~47.5) | 90 | | 32.5 (27~38.4) |
| **Housing** | | | | | | | | | | | | | | | | | |
| Live alone (owned or rented) | 45 | 64.3 (51.9~75.4) | 118 | | 57 (50~63.8) | 70 | | 57.4 (48.1~66.3) | | | | 93 | 60 (51.8~67.8) | | 163 | 58.8 (52.8~64.7) | |
| Live with someone else | 13 | 18.6 (10.3~29.7) | 49 | | 23.7 (18.1~30.1) | 24 | | 19.7 (13~27.8) | | | | 38 | 24.5 (18~32.1) | | 62 | 22.4 (17.6~27.8) | |
| Live at workplace or school | 8 | 11.4  (5.1~21.3) | 30 | | 14.5 (10~20) | 20 | | 16.4 (10.3~24.2) | | | | 18 | 11.6 (7~17.7) | | 38 | 13.7 (9.9~18.3) | |
| Other | 4 | 5.7 (1.6~14) | 10 | | 4.8 (2.3~8.7) | 8 | | 6.6 (2.9~12.5) | | | | 6 | 3.9 (1.4~8.2) | | 14 | 5.1 (2.8~8.3) | |
| **Marital status** | | | | | | | | | | | | | | | | | |
| Never married | 61 | 87.1 (77~93.9) | 182 | | 88.3 (83.2~92.4) | 105 | | 86.1 (78.6~91.7) | | | | 138 | 89.6 (83.7~93.9) | | 243 | 88 (83.6~91.6) | |
| Engaged or married | 5 | 7.1 (2.4~15.9) | 5 | | 2.4 (0.8~5.6) | 6 | | 4.9 (1.8~10.4) | | | | 4 | 2.6 (0.7~6.5) | | 10 | 3.6 (1.8~6.6) | |
| Separated or divorced | 4 | 5.7 (1.6~14) | 19 | | 9.2 (5.6~14) | 11 | | 9 (4.6~15.6) | | | | 12 | 7.8 (4.1~13.2) | | 23 | 8.3 (5.4~12.2) | |
| **Education** | | | | | | | | | | | | | | | | | |
| High school or below | 36 | 51.4 (39.2~63.6) | 90 | 43.5 (36.6~50.5) | | 58 | | 47.5 (38.4~56.8) | | | | 68 | 43.9 (35.9~52.1) | | 126 | 45.5 (39.5~51.6) | |
| Some college | 18 | 25.7 (16~37.6) | 50 | 24.2 (18.5~30.6) | | 26 | | 21.3 (14.4~29.6) | | | | 42 | 27.1 (20.3~34.8) | | 68 | 24.5 (19.6~30.1) | |
| College/Bachelors or above | 16 | 22.9 (13.7~34.4) | 67 | 32.4 (26~39.2) | | 38 | | 31.1 (23.1~40.2) | | | | 45 | 29 (22~36.9) | | 83 | 30 (24.6~35.7) | |
| **Annual income (USD)** | | | | | | | | | | | | | | | | | |
| Less than 3000 USD | 11 | 15.7 (8.1~26.4) | 64 | 30.9 (24.7~37.7) | | 38 | | 31.1 (23.1~40.2) | | | | 37 | 23.9 (17.4~31.4) | | 75 | 27.1 (21.9~32.7) | |
| Between 3001 ~ 6000 USD | 41 | 58.6 (46.2~70.2) | 88 | 42.5 (35.7~49.6) | | 60 | | 49.2 (40~58.4) | | | | 69 | 44.5 (36.5~52.7) | | 129 | 46.6 (40.6~52.6) | |
| Greater than 6000 USD | 18 | 25.7(16~37.6) | 55 | 26.6 (20.7~33.1) | | 24 | | 19.7 (13~27.8) | | | | 49 | 31.6 (24.4~39.6) | | 73 | 26.4 (21.3~32) | |

| **Table 2. Healthcare Services Uptake among 277 Transgender Individuals in China, 2020.** | |
| --- | --- |
| **Healthcare engagement** | **N (%)** |
| Hormone intervention history |  |
| -Have or currently undergoing | 118 (42.6) |
| -Never Used | 159(57.4) |
| Gender affirming surgery |  |
| -Have or currently undergoing | 26 (9.4) |
| -Never used | 251(90.6) |
| HIV testing* |  |
| -Yes | 220 (79.4) |
| -No | 57 (20.6) |
| STI testing in the past year^#^ |  |
| -Yes | 131(47.3) |
| -No | 146 (52.7) |
| PrEP use |  |
| -Yes | 24 (8.6) |
| -No | 252(90.7) |
| HIV-Positive | 20 (9.1) |
| Note: *Human Immuno-Deficiency Virus Testing;# Sexually Transmitted Infection Testing | |

| **Table 3. Factors correlated with gender identity disclosure to healthcareproviders among 277 transgender people in China , 2020.** | | | | | |
| --- | --- | --- | --- | --- | --- |
| **Variables** | | **Crude OR** | | **Adjusted OR*** | |
|  |  | **OR** | **95%CIa^#^** | **OR** | **95%CIs** |
| Ever had a stable sexual partner the past 3 months | No | Ref |  | Ref |  |
|  | Yes | 1.04 | 0.54-1.99 | 1.07 | 0.51-2.22 |
| Condom less sex | No | Ref |  | Ref |  |
|  | Yes | 0.84 | 0.20-3.04 | 0.81 | 0.16-4.13 |
| Ever tested for HIV | No | Ref |  | Ref |  |
|  | Yes | 1.85 | 1.03-3.35 | 1.72 | 0.87-3.39 |
| Ever tested for STI | No | Ref |  | Ref |  |
|  | Yes | 1.89 | 1.17-3.07 | 1.94 | 1.11-3.39 |
| Felt society is not fair to them | No | Ref |  | Ref |  |
|  | Yes | 0.95 | 0.59-1.53 | 0.99 | 0.58-1.67 |
| Had difficulties to get medical treatment | No | Ref |  | Ref |  |
|  | Yes | 0.81 | 0.50-1.30 | 0.73 | 0.43-1.23 |
| Rejected by family | No | Ref |  | Ref |  |
|  | Yes | 1.25 | 0.77-2.03 | 0.87 | 0..52-1.48 |
| Felt rejected due to gender | No | Ref |  | Ref |  |
|  | Yes | 1.43 | 0.87-2.34 | 1.39 | 0.79-2.42 |
| Felt do not get good medical services due to gender | No | Ref |  | Ref |  |
|  | Yes | 1.06 | 0.66-1.70 | 1.24 | 0.73-2.12 |
| Got emotional help and support from family | No | Ref |  | Ref |  |
|  | Yes | 0.96 | 0.59-1.56 | 0.98 | 0.58-1.67 |
| Ever smoked | No | Ref | 1.01-2.75 | Ref | 0.96-2.88 |
|  | Yes | 1.66 |  | 1.66 |  |
| Ever used alcohol | No | Ref |  | Ref |  |
|  | Yes | 1.74 | 1.07-2.84 | 1.16 | 0.66-2.04 |
| Ever used drugs | No | Ref |  | Ref |  |
|  | Yes | 1.52 | 0.73-3.29 | 1.18 | 0.50-2.78 |
| Felt comfortable disclosing gender identity to others | No | Ref |  | Ref |  |
|  | Yes | 1.08 | 0.67-1.74 | 1.28 | 0.75-2.18 |
| Hormone intervention history | No | Ref |  | Ref |  |
|  | Yes | 3.06 | 1.84-5.13 | 2.81 | 1.56-5.05 |
| Gender affirming surgery | No | Ref |  | Ref |  |
|  | Yes | 1.87 | 0.81-4.71 | 1.26 | 0.46-3.45 |
| PrEP use^§^ | No | Ref |  | Ref |  |
|  | Yes | 3.24 | 1.26-10.02 | 3.51 | 1.12-10.97 |
| Note: *Adjusted Odds Ratio;^#^95% Confidence Interval;^§^HIV Pre-Exposure Prophylaxis Use | | | | | |
