## Supplemental Data for "Disclosure of Gender Identity among Transgender Individuals to Healthcare Professionals in China: An Online Cross-sectional Study"

**Supplementary Table S1**

| **Behavior Characteristics Among Transgender Individuals 2020 (**n **= 277).** | | | | | | | | | | | | |
| --- | --- | --- | --- | --- | --- | --- | --- | --- | --- | --- | --- | --- |
|  | | **To Others** | | | | | **To Healthcare Professionals** | | | | **Overall (N = 277)** | |
|  |  | **Non-disclosers** | | **Disclosers** | | | **Non-disclosers** | | **Disclosers** | |  |  |
|  |  | **(n = 61, 22.0%)** | | **(n = 192, 78.0%)** | | | **(n = 122, 44.0%)** | | **(n = 155, 56.0%)** | |  |  |
|  |  | N | Percent (95%CI) | N | Percent (95%CI) | | N | Percent (95%CI) | N | Percent (95%CI) | N | Percent (95%CI) |
| **Lifetime sex work involvement** | | | | | | | | | | | | |
|  | Yes | 29 | 41.4 (29.8~53.8) | 87 | | 42.4 (35.6~49.5) | 47 | 38.8 (30.1~48.1) | 69 | 44.8 (36.8~53) | 116 | 42.2 (36.3~48.3) |
|  | No | 41 | 58.6 (46.2~70.2) | 118 | | 57.6 (50.5~64.4) | 74 | 61.2 (51.9~69.9) | 85 | 55.2 (47~63.2) | 159 | 57.8 (51.7~63.7) |
| **Ever had a stable sexual partner the past 3 months** | | | | | | | | | | | | |
|  | Yes | 18 | 52.9 (35.1~70.2) | 62 | | 53 (43.5~62.3) | 32 | 52.5 (39.3~65.4) | 48 | 53.3 (42.5~63.9) | 80 | 53 (44.7~61.1) |
|  | No | 16 | 47.1 (29.8~64.9) | 55 | | 47 (37.7~56.5) | 29 | 47.5 (34.6~60.7) | 42 | 46.7 (36.1~57.5) | 71 | 47 (38.9~55.3) |
| **Condom Less Sex** | | | | | | | | | | | | |
|  | Yes | 16 | 88.9 (65.3~98.6) | 53 | | 85.5 (74.2~93.1) | 4 | 12.5 (3.5~29) | 7 | 14.6 (6.1~27.8) | 11 | 13.8 (7.1~23.3) |
|  | No | 2 | 11.1 (1.4~34.7) | 9 | | 14.5 (6.9~25.8) | 7 | 21.9 (9.3~40) | 8 | 16.7 (7.5~30.2) | 15 | 18.8 (10.9~29) |
| **Felt society is not fair to them** | | | | | | | | | | | | |
|  | Disagree | 39 | 55.7 (43.3~67.6) | 90 | | 43.5 (36.6~50.5) | 56 | 45.9 (36.8~55.2) | 73 | 47.1 (39~55.3) | 129 | 46.6 (40.6~52.6) |
|  | Agree | 31 | 44.3 (32.4~56.7) | 117 | | 56.5 (49.5~63.4) | 66 | 54.1 (44.8~63.2) | 82 | 52.9 (44.7~61) | 148 | 53.4 (47.4~59.4) |
| **Had difficult  to getting medical treatment** | | | | | | | | | | | | |
|  | Yes | 36 | 51.4 (39.2~63.6) | 108 | | 52.2 (45.1~59.1) | 67 | 54.9 (45.7~63.9) | 77 | 49.7 (41.6~57.8) | 144 | 52 (45.9~58) |
|  | No | 34 | 48.6 (36.4~60.8) | 99 | | 47.8 (40.9~54.9) | 55 | 45.1 (36.1~54.3) | 78 | 50.3 (42.2~58.4) | 133 | 48 (42~54.1) |
| **Rejected by family** | | | | | | | | | | | | |
|  | Yes | 27 | 38.6 (27.2~51) | 90 | | 43.7 (36.8~50.8) | 48 | 39.3 (30.6~48.6) | 69 | 44.8 (36.8~53) | 117 | 42.4 (36.5~48.5) |
|  | No | 43 | 61.4 (49~72.8) | 116 | | 56.3 (49.2~63.2) | 74 | 60.7 (51.4~69.4) | 85 | 55.2 (47~63.2) | 159 | 57.6 (51.5~63.5) |
| **Felt rejected due to gender** | | | | | | | | | | | | |
|  | Disagree | 47 | 67.1 (54.9~77.9) | 124 | | 59.9 (52.9~66.6) | 81 | 66.4 (57.3~74.7) | 90 | 58.1 (49.9~65.9) | 171 | 61.7 (55.7~67.5) |
|  | Agree | 23 | 32.9 (22.1~45.1) | 83 | | 40.1 (33.4~47.1) | 41 | 33.6 (25.3~42.7) | 65 | 41.9 (34.1~50.1) | 106 | 38.3 (32.5~44.3) |
| **Felt do not get Good Medical services due to gender** | | | | | | | | | | | | |
|  | Disagree | 32 | 45.7 (33.7~58.1) | 93 | | 44.9 (38~52) | 56 | 45.9 (36.8~55.2) | 69 | 44.5 (36.5~52.7) | 125 | 45.1 (39.2~51.2) |
|  | Agree | 38 | 54.3 (41.9~66.3) | 114 | | 55.1 (48~62) | 66 | 54.1 (44.8~63.2) | 86 | 55.5 (47.3~63.5) | 152 | 54.9 (48.8~60.8) |
| **Got emotional help and support from family** | | | | | | | | | | | | |
|  | Disagree | 22 | 31.4 (20.9~43.6) | 84 | | 40.6 (33.8~47.6) | 46 | 37.7 (29.1~46.9) | 60 | 38.7 (31~46.9) | 106 | 38.3 (32.5~44.3) |
|  | Agree | 48 | 68.6 (56.4~79.1) | 123 | | 59.4 (52.4~66.2) | 76 | 62.3 (53.1~70.9) | 95 | 61.3 (53.1~69) | 171 | 61.7 (55.7~67.5) |
| **Ever smoked** | | | | | | | | | | | | |
|  | Yes | 23 | 32.9 (22.1~45.1) | 79 | | 38.2 (31.5~45.2) | 37 | 30.3 (22.3~39.3) | 65 | 41.9 (34.1~50.1) | 102 | 36.8 (31.1~42.8) |
|  | No | 47 | 67.1 (54.9~77.9) | 128 | | 61.8 (54.8~68.5) | 85 | 69.7 (60.7~77.7) | 90 | 58.1 (49.9~65.9) | 175 | 63.2 (57.2~68.9) |
| **Ever used alcohol** | | | | | | | | | | | | |
|  | Yes | 33 | 47.1 (35.1~59.4) | 135 | | 65.2 (58.3~71.7) | 65 | 53.3 (44~62.4) | 103 | 66.5 (58.4~73.8) | 168 | 60.6 (54.6~66.4) |
|  | No | 37 | 52.9 (40.6~64.9) | 72 | | 34.8 (28.3~41.7) | 57 | 46.7 (37.6~56) | 52 | 33.5 (26.2~41.6) | 109 | 39.4 (33.6~45.4) |
| **Ever used drugs** | | | | | | | | | | | | |
|  | Yes | 33 | 47.1 (35.1~59.4) | 135 | | 65.2 (58.3~71.7) | 12 | 9.8 (5.2~16.6) | 22 | 14.2 (9.1~20.7) | 34 | 12.3 (8.7~16.7) |
|  | No | 37 | 52.9 (40.6~64.9) | 72 | | 34.8 (28.3~41.7) | 110 | 90.2 (83.4~94.8) | 133 | 85.8 (79.3~90.9) | 243 | 87.7 (83.3~91.3) |
| **Felt comfortable disclosing gender identity to others** | | | | | | | | | | | | |
|  | Disagree | 33 | 47.1 (35.1~59.4) | 135 | | 65.2 (58.3~71.7) | 59 | 48.4 (39.2~57.6) | 72 | 46.5 (38.4~54.6) | 131 | 47.3 (41.3~53.4) |
|  | Agree | 37 | 52.9 (40.6~64.9) | 72 | | 34.8 (28.3~41.7) | 63 | 51.6 (42.4~60.8) | 83 | 53.5 (45.4~61.6) | 146 | 52.7 (46.6~58.7) |

**Supplementary Table S2**

| **Factors Correlated with Gender Identity Disclosure** | | | | | |
| --- | --- | --- | --- | --- | --- |
| **Variables** | | **Crude Model** | | **Adjusted OR^[1]^** | |
|  |  | **OR** | **95%C.Is** | **OR** | **95%C.Is^[2]^** |
| Ever had a stable sexual partner in the past 3 months | No | *Ref* | 0.46-2.16 | *Ref* | 0.58-3.61 |
|  | Yes | 1.00 |  | 1.45 |  |
| Condom less Sex | No | *Ref* | 0.11-3.24 | *Ref* | 0.16-6.26 |
|  | Yes | 0.74 |  | 1.02 |  |
| Ever had anal or vaginal sex with sex partner | No  Yes | *Ref*  1.13 | 0.25-6.04 | *Ref*  0.35 | 0.05-2.32 |
| Ever tested for HIV^[3]^ | No  Yes | *Ref*  1.49 | 0.77-2.79 | *Ref*  1.87 | 0.77-4.54 |
| Ever tested for STI^[4]^ | No  Yes | *Ref*  1.74 | 1.01-3.07 | *Ref*  1.82 | 0.98-3.39 |
| Felt society is not fair to them | No  Yes | *Ref*  1.64 | 0.95-2.84 | *Ref*  1.45 | 0.80-2.63 |
| Had difficulties to get Medical treatment | No  Yes | *Ref*  1.03 | 0.59-1.77 | *Ref*  0.83 | 0.44-1.55 |
| Rejected by family | No  Yes | *Ref*  1.24 | 0.71-2.17 | *Ref*  0.93 | 0.50-1.71 |
| Felt rejected due to gender | No  Yes | *Ref*  1.37 | 0.78-2.45 | *Ref*  0.70 | 0.37-1.32 |
| Felt do not get Good Medical services due to gender | No  Yes | *Ref*  1.03 | 0.59-1.78 | *Ref*  1.19 | 0.65-2.19 |
| Got emotional help and support from family | No  Yes | *Ref*  0.67 | 0.37-1.18 | *Ref*  0.87 | 0.47-1.63 |
| Ever smoked | No  Yes | *Ref*  1.26 | 0.72-2.26 | *Ref*  0.76 | 0.41-1.44 |
| Ever used alcohol | No  Yes | *Ref*  2.10 | 1.22-3.66 | *Ref*  2.44 | 1.312-4.55 |
| Ever used drugs | No  Yes | *Ref*  2.79 | 1.05-9.68 | *Ref*  0.37 | 0.12-1.12 |
| Felt comfortable disclosing gender identity to others | No  Yes | *Ref*  1.07 | 0.62-1.84 | *Ref*  1.48 | 0.79-2.76 |
| Hormone intervention history | No  Yes | *Ref*  2.06 | 1.17-3.73 | *Ref*  1.86 | 0.93-3.74 |
| Ever had gender affirming surgery | No  Yes | *Ref*  0.91 | 0.38-2.42 | *Ref*  0.68 | 0.23-2.06 |
| Ever used PrEP^[5]^ | No  Yes | *Ref*  2.48 | 0.82-10.76 | *Ref*  3.04 | 0.78-11.89 |
| Note:2= 95% Confidence Intervals 3= Human Immuno-Deficiency Virus;4= Sexually Transmitted Infections; 5= Pre-Exposure Prophylaxis Use | | | | | |

1. Note: Odds Ratio
2. 95% Confiden
